# Association of Alzheimer’s disease polygenic risk score with concussion severity and recovery metrics

**DOI:** 10.1101/2024.07.10.24309042

**Authors:** Kaitlyn M. Dybing, Thomas W. McAllister, Yu-Chien Wu, Brenna C. McDonald, Steven P. Broglio, Jason P. Mihalik, Kevin M. Guskiewicz, Joshua T. Goldman, Jonathan C. Jackson, Shannon L. Risacher, Andrew J. Saykin, Kelly N. H. Nudelman

**Author notes:** co-corresponding authors: Kaitlyn M. Dybing., Shannon L. Risacher, Kelly N. H. Nudelman.

## Abstract

Identification of genetic alleles associated with both Alzheimer’s disease (AD) and concussion severity/recovery could help explain the association between concussion and elevated dementia risk. However, there has been little investigation into whether AD risk genes associate with concussion severity/recovery, and the limited findings are mixed. We used AD polygenic risk scores (PRS) and *APOE* genotypes to investigate any such associations in the NCAA-DoD Grand Alliance CARE Consortium (CARE) dataset. We assessed six outcomes in 931 total participants. The outcomes were two concussion recovery measures (number of days to asymptomatic status, number of days to return to play (RTP)) and four concussion severity measures (scores on SAC and BESS, SCAT symptom severity, and total number of symptoms). We calculated PRS using a published score [1] and performed multiple linear regression (MLR) to assess the relationship of PRS with the outcomes. We also used t-tests and chi-square tests to examine outcomes by *APOE* genotype, and MLR to analyze outcomes in European and African genetic ancestry subgroups. Higher PRS was associated with longer injury to RTP in the normal RTP (<24 days) subgroup (*p* = 0.024), and one standard deviation increase in PRS resulted in a 9.89 hour increase to the RTP interval. There were no other consistently significant effects, suggesting that high AD genetic risk is not strongly associated with more severe concussions or poor recovery in young adults. Future studies should attempt to replicate these findings in larger samples with longer follow-up using PRS calculated from diverse populations.

## Introduction

Sport-related concussions are a serious public health concern, with an annual occurrence of 1.6 to 3.8 million in the United States [2–4]. Outside of acute consequences, such as inability to participate in athletic competition and/or academic difficulties [5–8], concussions can be associated with long-term consequences, especially when multiple injuries are incurred [6, 9–14]. Concussion has also been linked to elevated risk for neurodegenerative diseases like chronic traumatic encephalopathy (CTE) [15–20]. However, there have been limited opportunities to examine associations between concussion incurred during the early decades of life and later dementia risk, as longitudinal clinical studies can be costly and challenging [21, 22]. Additionally, dementia research cohorts are often overwhelmingly white and highly educated [23, 24], making it difficult to investigate diverse populations. The Concussion Assessment, Research and Education (CARE) Consortium was designed to address many of these limitations, and its mission is to expand and improve concussion diagnosis, treatment, and prevention [25]. The CARE dataset encompasses both non-military NCAA collegiate athletes and Military Service Academy students and Military Service Academy NCAA-student athletes and is ethnically and racially diverse [25].

Previous reports have suggested concussion severity and/or recovery may be influenced by genetic factors. Specifically, the ε4 allele of the Apolipoprotein E (*APOE*) gene, also known as *APOE* ε4, is associated with higher likelihood of unfavorable outcome, such as reduced cognitive functioning, after concussion [26–33]. Importantly, *APOE* ε4 also contributes to elevated risk for Alzheimer’s disease (AD) [34], a progressive neurodegenerative disorder that clinically presents with loss of memory, cognitive function, and behavioral changes. AD is pathologically characterized by extracellular amyloid-β plaques, intracellular neurofibrillary tau tangles, and neurodegeneration [35, 36]. In addition to *APOE* ε4, a myriad of other genetic variants contribute to risk for AD [37]. The overall disease risk of an individual genomic profile can be summarized as a polygenic risk score (PRS), a weighted sum of the risk alleles present in an individual [38–40]. PRS are widely used in both genetic and neurodegenerative disease research [38–40].

The previously described association of *APOE* ε4 with elevated risk for both AD and poor concussion prognosis raises the question of whether additional genetic links may exist. However, previous research has concentrated on *APOE* ε4 [26–32], and other genes are under-investigated. Therefore, we sought to characterize associations of AD PRS and *APOE* genotype with concussion severity and recovery metrics to assess whether young adult individuals with high genetic risk for AD would experience poorer concussion recovery and/or more severe injuries.

## Methods

### Study sample

This study utilized data collected from participants in the multi-site Concussion Assessment, Research, and Education (CARE) Consortium established by the National Collegiate Athletic Association (NCAA) and US Department of Defense (DoD), protocols of which have been described in previous reports [25]. Briefly, NCAA student athletes and Military Service Academy students recruited from participating institutions complete a baseline test battery incorporating demographics, medical history, cognitive performance, and other variables. If at any time point a participant is suspected of having suffered a concussion, evaluation and diagnosis are made by on-site medical personnel. Injured participants are then assessed at five timepoints: within 6 hours of the injury, again at 24-48 hours post-injury, once asymptomatic and cleared to initiate return to play protocols, when fully cleared to return to play, and finally approximately 6 months after the injury. Until participants are cleared to return to activity, symptoms are documented daily for up to 14 days then once weekly thereafter. Tests conducted at each post-injury timepoint include the Sport Concussion Assessment Tool (SCAT) for symptomology and symptom severity, the Standardized Assessment of Concussion (SAC) to assess cognitive performance, the Balance Error Scoring System as a measure of postural stability, and many others.

To date, the CARE Consortium has enrolled over 20,000 participants. However, not all incurred an injury while in the study. Upon initiation of this project, there were 1,917 recorded concussions in the dataset. Of these, 304 injuries did not have associated outcome measures and were removed. Furthermore, 573 concussions did not have corresponding genetic data and were excluded. Also, only considered participants’ first injury in the study was considered, thus, 107 repeat injuries were removed due to concern over the introduction of bias on the outcomes and bias due to multiple instances of the same PRS.

We also wanted to remove first- or second-degree relatives from the sample. To do so, we employed the pi-hat identical-by-descent (IBD) estimate from the PLINK software package [41]. Higher pi-hat values represent greater genetic similarity, and we used a threshold of 0.2 to indicate first- or second-degree relatives [41]. In our sample, two pairs of participants had pi-hat > 0.2, and one from each pair was randomly removed. This left 931 non-related injured participants for analysis. These participants were further subdivided into four groups based on sex and whether their concussion was associated with loss of consciousness (LOC): females with loss of consciousness (F LOC+), females without loss of consciousness (F LOC-), males with loss of consciousness (M LOC+), and males without loss of consciousness (M LOC-) (Figure 1). A consort diagram of the participant selection process is available as Figure 1.

**Fig. 1.**
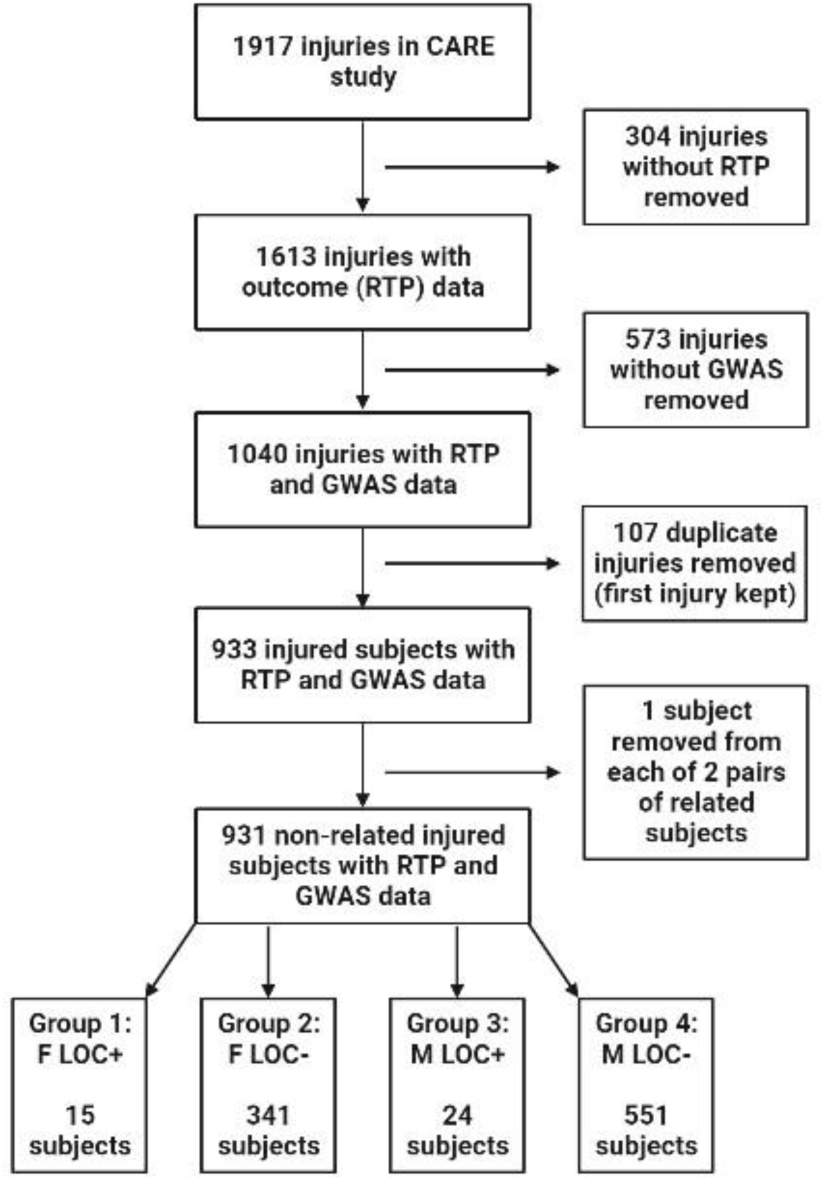
A consort diagram detailing the cohort of CARE participants used in this study (created with BioRender.com)

### Determining polygenic risk score and APOE genotype

To calculate AD PRS, we selected a published score [1] comprised of 39 single-nucleotide polymorphisms (SNPs) and used it to calculate PRS in our participants. We selected this score principally because of the large sample size (over 400,000) from which it was derived [1]. Once calculated, PRS were transformed to z-scores. The CARE data was missing 2 alleles from the original PRS: rs2732703 (a duplicated variant with different weights for *APOE* ε4 carriers and non-carriers) and rs616338. We therefore calculated the PRS from 37 SNPs in our participants. There were several instances where a participant did not have available data for one or more SNP(s). There are two analytical strategies for addressing this: the missing SNP can be ignored and the PRS calculated without it, or the value of the missing SNP can be substituted with the mean from all other participants on that SNP. To compare these methods, we performed a sensitivity analysis and did not identify significant differences between these strategies (*Supplemental figures 1a-d*). We therefore chose to use the ignored missing values strategy for all further analyses.

We also wanted to investigate recovery/severity outcomes as a function of *APOE* genotype. As we had limited power, we split participants into *APOE* ε3/ε3, ε2 carriers, and ε4 carriers rather than considering every possible *APOE* genotype. Due to limited power, we chose not to further stratify participants. Of the 931 participants, 10 did not have available *APOE* genotypes and 21 were *APOE* ε2/ε4. The ε2/ε4 participants were excluded given the small number and potential for confounding if included in either the ε2 or ε4 carrier group. Therefore, 900 total participants were included.

### Selection of outcome metrics

There is a significant amount of data concerning both concussion severity and recovery generated through the CARE Consortium [25]; we examined six key metrics. These were the injury to asymptomatic interval, the injury to RTP interval, SCAT symptom severity and total symptom scores, and total scores on the SAC and BESS.

First, the injury to return to play (RTP) interval, also called days to RTP or simply RTP, represents the length of time an individual takes to be medically cleared for full participation in sports/physical activity after a concussion [42, 43]. This is thought to typically occur within one month of injury [6, 44–49], but may vary depending on factors like age [50, 51] and sex [52–55]. Athletes enrolled in the CARE Consortium initiated the RTP protocol at the discretion of staff and were not necessarily entirely asymptomatic when the protocol was initiated [56]. Though the international consensus group on concussion in sport defined normal recovery to be <28 days [6], a previous report from the CARE Consortium suggested 80% of CARE participants RTP within 24 days [43]. Therefore, we used <24 days as the definition of normal RTP in this study. We also considered the injury to asymptomatic interval, which tracks how long a participant remains symptomatic after an injury [43, 49]. In previous studies by the CARE team, 80% of participants were found to be asymptomatic by day 14 after concussion [43].

The Sport Concussion Assessment Tool (5th edition) (SCAT5) is one of the most commonly used tests to evaluate concussion. The SCAT5 incorporates elements such as reading, memory, balance, and gait [57]. Individuals self-report 22 concussion symptoms on a 7-point Likert scale, ranging from 0 (none) to 6 (severe) [57, 58]. This generates a score out of 22 reflecting the total number of symptoms experienced by the individual (SCAT5 total symptoms score) and a score out of 132 indicating the severity of the present symptoms (SCAT5 symptom severity score) [58]. The SCAT5 incorporates the BESS and SAC (described below); from this test we utilized only the SCAT5 symptom severity and total symptoms scores.

Another validated and widely used test for assessing concussions is the Balance Error Scoring System (BESS), which consists of 3 stances performed on both a firm and a foam surface with the eyes closed [59]. Participants are scored from 0 to 30 based on the number of errors made within 20-second trials, with higher scores indicating greater postural instability [59]. Errors include opening the eyes or falling out of the stance, among others [59]. The BESS can identify balance deficits/postural instability in concussed individuals [60–62].

Finally, the Standardized Assessment of Concussion (SAC) is a brief screening tool to assess cognition in suspected concussion through four domains (orientation, immediate memory, concentration, and delayed recall) [63]. Participants receive a total score out of 30, where lower scores indicate poorer performance [63].

### Statistical analyses

Statistical tests were performed using RStudio 2023.12.0 and SPSS 29.0.1.0. A *p*-value < 0.05 was considered significant for all comparisons. Demographic comparisons were performed in SPSS using chi-square tests or one-way ANOVAs with Tukey post-hoc tests. Chi-square tests were used to determine the expected number of participants in each category in comparison to the actual number of participants in each category. If the homogeneity of variance assumption was violated (indicated by a significant Levene statistic), the Welch test was used in place of ANOVA and post-hoc tests were performed using the Games-Howell method. When comparing AD PRS with outcomes, we performed linear regressions using the lm() function in R. When comparing outcomes by *APOE* genotype, t-tests were performed using the t_test() function in R.

## Results

### Demographic & neuropsychological variables

Participant characteristics are summarized in Table 1. We analyzed participants’ baseline demographics and neuropsychological characteristics using the first available test values obtained within 24-48 hours following the injury (Table 1). F LOC- participants were younger (19.85 years) than M LOC- participants (20.20 years) (mean difference = -0.347 years, *p* < 0.001, 95% CI [-0.572, - 0.121]). Additionally, F LOC- participants had higher SCAT total symptoms scores (10.28) than M LOC+ participants (5.33) (mean difference = 4.942, *p* = 0.028, η2 = 0.018, 95% CI [0.385, 9.50]). Furthermore, F LOC- participants took longer to become asymptomatic (12.52 days) than M LOC- participants (9.54 days) (mean difference 2.98, *p* = 0.014, 95% CI [0.431, 5.530).

**Table 1.** Baseline demographic & neuropsychological characteristics (at first available post-injury assessment) of included participants.

|  | F LOC+ | F LOC- | M LOC+ | M LOC- | N (%) | Levene statistic (p) | ANOVA F/Welch statistic | P-value |
| --- | --- | --- | --- | --- | --- | --- | --- | --- |
| Number of participants | 15 | 341 | 24 | 551 | 931 | - | - | - |
| Military participants (%) | 0 (0) | 70 (20.5) | 12 (50) | 176 (31.9) | 258 (27.7) | - | - | - |
| Civilian participants (%) | 15 (100) | 271 (79.5) | 12 (50) | 375 (68.1) | 673 (72.3) | - | - | - |
| Avg. AD PRS z-score (SD) | -0.073 (0.54) | -0.0044 (1.03) | 0.045 (1.11) | 0.0028 (0.99) | 931 | 2.006 (.112) | .046 | 0.987 |
| *Sport* |  |  |  |  |  |  |  |  |
| Baseball | - | - | 1 | 21 | 22 |  |  |  |
| Basketball | 1 | 35 | - | 18 | 54 |  |  |  |
| Beach volleyball | - | 1 | - | - | 1 |  |  |  |
| Boxing | - | - | - | 2 | 2 |  |  |  |
| Cheerleading | - | 17 | - | 4 | 21 |  |  |  |
| Cross country/track | - | 6 | - | 31 | 37 |  |  |  |
| Diving | - | 11 | - | 6 | 17 |  |  |  |
| Fencing | - | 2 | - | 1 | 3 |  |  |  |
| Field event | - | 6 | - | 2 | 8 |  |  |  |
| Field hockey | 1 | 16 | - | - | 17 |  |  |  |
| Football | - | - | 5 | 262 | 267 |  |  |  |
| Golf | - | 2 | - | - | 2 |  |  |  |
| Gymnastics | 3 | 12 | - | 5 | 20 |  |  |  |
| Ice hockey | - |  | - | 15 | 15 |  |  |  |
| Lacrosse | - | 24 | 2 | 29 | 55 |  |  |  |
| Rowing/crew | - | 7 | - | - | 7 |  |  |  |
| Rugby | - | 11 | 2 | 13 | 26 |  |  |  |
| Soccer | 7 | 59 | 5 | 50 | 121 |  |  |  |
| Softball | - | 23 | - | - | 23 |  |  |  |
| Sprint football | - | - | - | 1 | 1 |  |  |  |
| Swimming | - | 12 | - | 7 | 19 |  |  |  |
| Tennis | - | 5 | 1 | 4 | 10 |  |  |  |
| Track/field | 3 | - | - | - | 3 |  |  |  |
| Volleyball | - | 39 | - | 5 | 44 |  |  |  |
| Water polo | - | 18 | - | 8 | 26 |  |  |  |
| Wrestling | - | - | 1 | 22 | 23 |  |  |  |
| Blank/military student | - | 35 | 7 | 45 | 87 |  |  |  |
| APOE genotype |  |  |  |  |  |  |  |  |
| ε3/ε3 | 9 | 206 | 13 | 320 | 548<br>(58.9) | χ<sup>2</sup> (12, N = 931) = 17.95, *p* = 0.117 |  |  |
| ε2 carrier (ε2/ε2 or ε2/ε3) | 2 | 44 | 2 | 61 | 109<br>(11.7) |  |  |  |
| ε4 carrier (ε3/ε4 or ε4/ε4) | 4 | 89 | 8 | 142 | 243<br>(26.1) |  |  |  |
| ε2/ε4 (excluded) | 0 | 1 | 0 | 20 | 21<br>(2.26) |  |  |  |
| Missing/no data | 0 | 1 | 1 | 8 | 10<br>(1.07) |  |  |  |
| Self-reported race/ethnicity (%) |  |  |  |  |  |  |  |  |
| Non-Hispanic white | 10 (66.7) | 213<br>(62.5) | 17 (70.8) | 320<br>(59.18) | 560<br>(60.15) | χ<sup>2</sup> (21, N = 931) = 36.064, *p* = **0.022**<br>*F LOC- Black/African American*<br>*(expected 49)*<br>*M LOC- Black/African American*<br>*(expected 79)* |  |  |
| Black/African American | 2 (13.3) | **23 (6.7)** | 4 (16.7) | **104**<br>**(18.9)** | 133<br>(14.3) |  |  |  |
| Hispanic white | 1 (6.7) | 14 (4.1) | 0 | 18 (3.27) | 33<br>(3.54) |  |  |  |
| Asian | 0 (0) | 8 (2.3) | 0 | 9 (1.8) | 17<br>(1.83) |  |  |  |
| Multiple races | 2 (13.3) | 34 (10.0) | 0 | 39 (7.1) | 75<br>(8.06) |  |  |  |
| Hawaiian/Pacific Islander | 0 | 3 (0.9) | 0 | 2 (0.4) | 5 (0.54) |  |  |  |
| Native American/Indian/Alaskan | 0 | 0 | 0 | 1 (0.2) | 1 (0.11) |  |  |  |
| Skipped | 0 | 46 (13.5) | 3 (12.5) | 58<br>(10.53) | 107<br>(11.49) |  |  |  |
| Genetic ancestry |  |  |  |  |  |  |  |  |
| European/EUR | 12 | **280** | 20 | **419** | 731<br>(78.52) | χ<sup>2</sup> (3, N = 866) = 16.067, *p* = **0.001**<br>*F LOC- (expected 261 EUR, 48 AFR)*<br>*M LOC- (expected 440 EUR, 81 AFR)* |  |  |
| African/AFR | 1 | **29** | 3 | **102** | 135<br>(14.50) |  |  |  |
| Outcomes |  |  |  |  |  |  |  |  |
| Avg. age at injury (SD) | 20.35<br>(1.62) | 19.85<br>(1.12) | 19.84<br>(1.02) | 20.20<br>(1.35) | 931 | 4.866<br>**(0.002)** | 6.026<sup>a</sup> | **0.002<sup>a</sup>**<br>(F LOC- vs. M LOC-; *p* < **.001**) |
| Avg. days to RTP after injury (SD) | 17.5<br>(10.46) | 23.63<br>(23.48) | 20.04<br>(21.63) | 20.27<br>(26.11) | 931 | 1.478<br>(0.219) | 1.433 | 0.232 |
| Avg. days to asymptomatic after injury (SD) | 9.74<br>(5.26) | 12.52<br>(16.50) | 10.36<br>(20.36) | 9.54<br>(7.81) | 857 | **7.689**<br>**(<0.001)** | 2.938<sup>a</sup> | **0.045<sup>a</sup>** (F LOC- vs M LOC-; *p* = **0.014**). |
| Avg. SCAT # of symptoms (SD) (*n participants*) | 9.44<br>(4.48) (9) | 10.28<br>(5.94) (225) | 5.33<br>(6.53) (12) | 9.61<br>(6.01) (225) | 471 | 0.672<br>(0.569) | 2.805 | **0.039**<br>(F LOC- vs M LOC+; *p* = **.028**) |
| Avg. SCAT symptom severity score (SD) (*n participants*) | 19.33<br>(12.71) (9) | 22.91<br>(20.24) (225) | 10.67<br>(17.20) (12) | 20.61<br>(19.42) (225) | 471 | 1.201<br>(0.309) | 1.784 | 0.149 |
| Avg. BESS total score<br>(SD) (*n participants*) | 12.27<br>(5.56)<br>(15) | 14.33<br>(7.71)<br>(323) | 13.96<br>(7.60)<br>(23) | 14.77<br>(7.45)<br>(527) | 888 | 0.682<br>(0.563) | 0.744 | 0.526 |
| Avg. SAC total score<br>(SD) (*n participants*) | 25.07<br>(6.73)<br>(15) | 26.95<br>(2.08)<br>(336) | 26.71<br>(2.16)<br>(24) | 26.76<br>(2.40)<br>(543) | 918 | 12.628<br>(**<0.001**) | 0.866<sup>a</sup> | 0.466 |

<sup>a</sup> Welch statistic & Welch test p-value reported due to significant Levene statistic

The chi-square test of self-reported ancestry was significant (χ2 (21, N = 931) = 36.064, *p* = 0.022, Cramer’s V = 0.114). There were fewer Black/African American F LOC- participants and more Black/African American M LOC- participants than anticipated. The chi-square test of genetic ancestry was also significant (χ2 (3, N = 866) = 16.067, *p* = 0.001, Cramer’s V = 0.136). In the F LOC- group, there were more European participants but fewer African participants than anticipated. Conversely, in the M LOC- group, there were fewer European participants but more African participants than anticipated. The chi-square test of *APOE* genotype was not significant (χ2 (12, N = 931) = 17.95, *p* = 0.117), and there were no additional differences in the outcomes.

### Concussion recovery (injury to RTP and injury to asymptomatic intervals) as a function of PRS

We assessed the number of days a participant took to return to play (injury to RTP interval/RTP) by AD PRS using linear regression (Table 2). We divided the participants into two categories: normal (<24 days) and long (>24 days) RTP [43]. We found a relationship between PRS and RTP in the normal RTP category, where higher PRS was associated with a longer injury to RTP interval (β = 0.412, SE = 0.182, 95% CI [0.055, 0.769], t-value = 2.267, *p* = 0.024) (Figure 2a). There was no relationship in the long RTP category (*p* = 0.778) (Figure 2b).

**Fig. 2.**
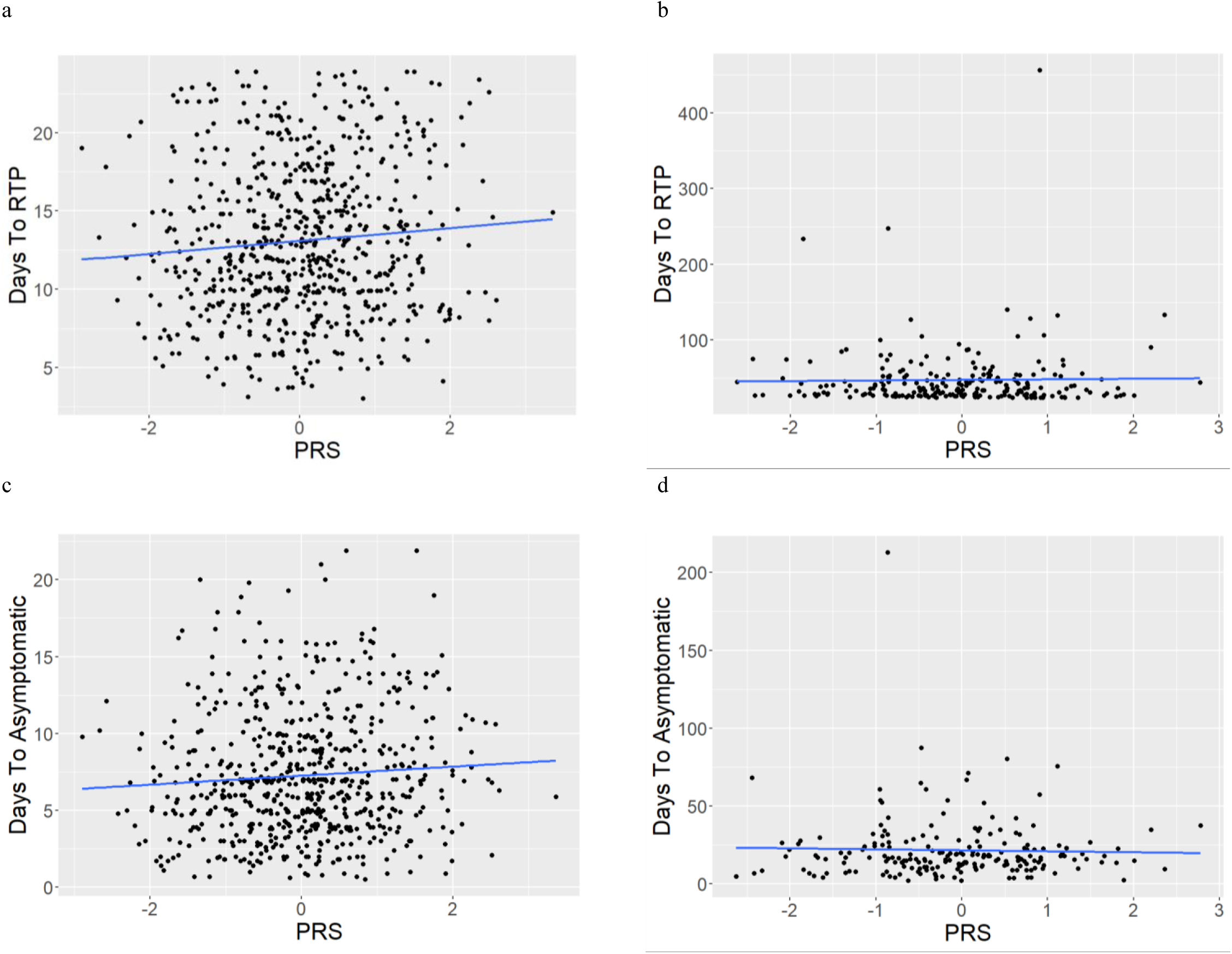
Concussion recovery measures as a function of PRS. Number of days to RTP by PRS in normal (<24 days) (a) and long (>24 days) (b) RTP categories, and number of days to asymptomatic in normal RTP participants (c) and long RTP participants (d)

**Table 2.**
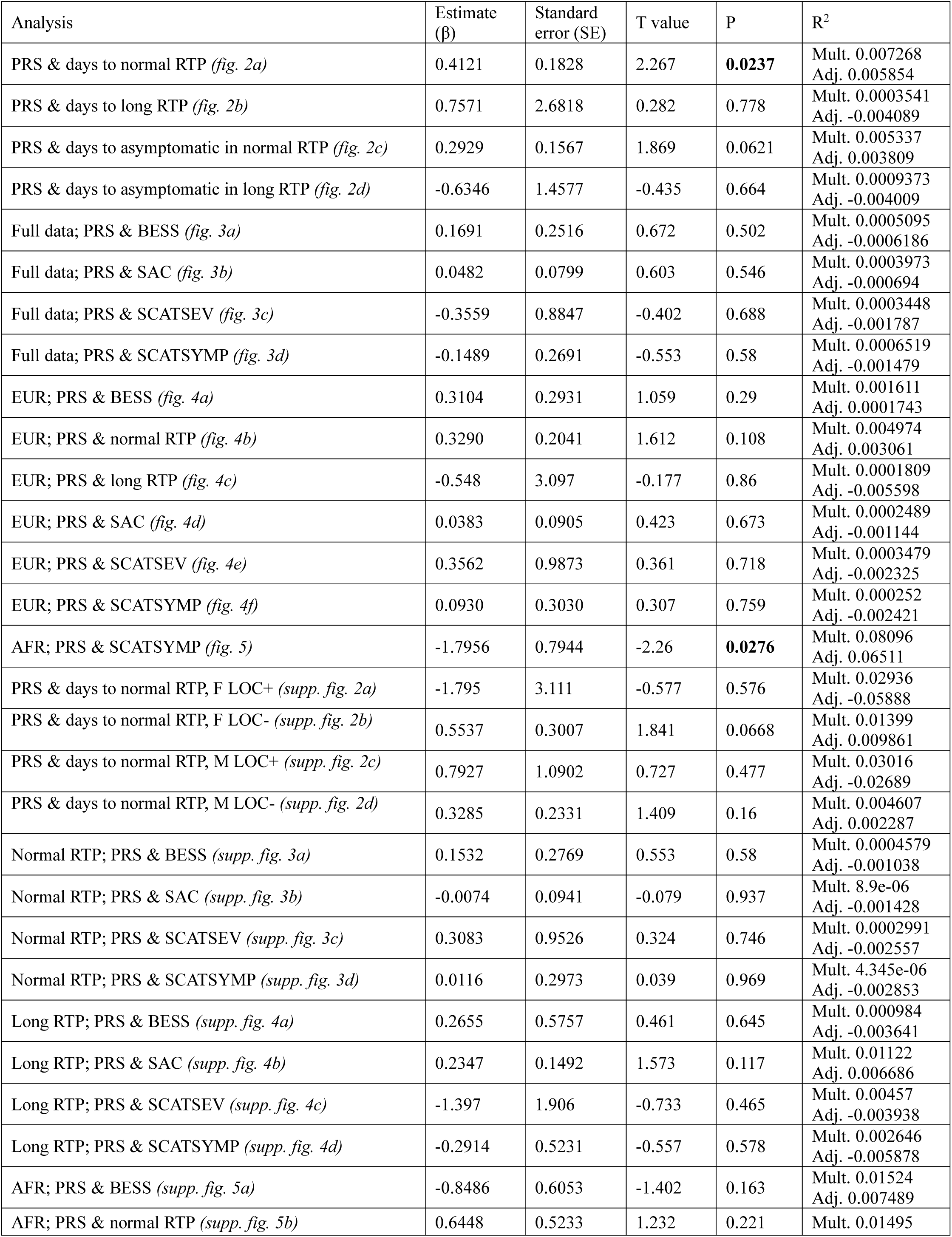
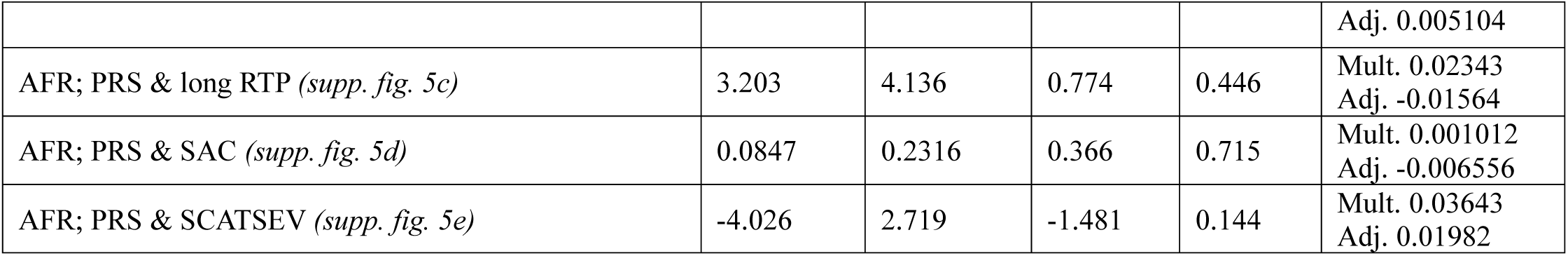
Summary of outcomes of linear regression analyses.

Since there was a significant relationship between PRS and RTP in the whole normal RTP group, we analyzed the injury to RTP interval by PRS in each participant subgroup *(supplementary figures 2a-d).* There was no relationship in the F LOC+ group (*p* = 0.576), the M LOC+ group (*p* = 0.477), or the M LOC- group (*p* = 0.16), and the F LOC- group approached significance (*p* = 0.067) (Table 2).

We then assessed the injury to asymptomatic interval as a function of PRS with participants split into normal and long RTP subsets (Table 2). This was done rather than splitting based on the normal injury to asymptomatic interval (<14 days, as defined previously [43]) so that the results could be directly compared with the RTP analysis. The relationship approached significance in the normal RTP group (*p* = 0.062) (Figure 2c) but was not significant in the long RTP group (*p* = 0.664) (Figure 2d).

### Concussion severity outcomes as a function of PRS

Linear regressions were used to assess the impact of AD PRS on concussion severity outcomes (BESS and SAC total scores, SCAT 5 symptom severity score (SCATSEV) and total number of symptoms (SCATSYMP)) (Table 2). No significant relationships were identified in the full data (BESS *p* = 0.502; SAC *p* = 0.546; SCATSEV *p* = 0.688; SCATSYMP *p* = 0.58) (Figures 3a-d).

**Fig. 3.**
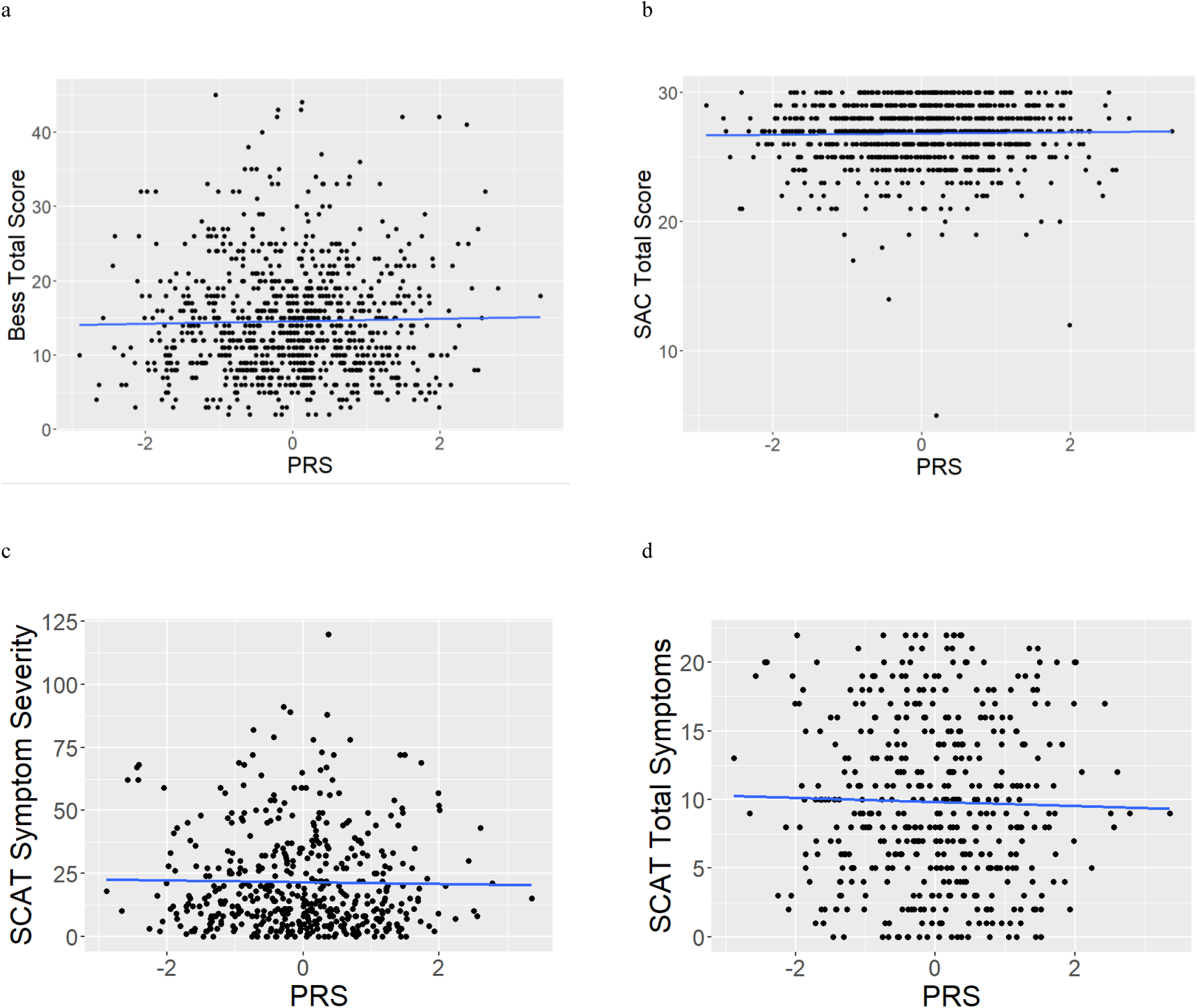
AD PRS and severity outcome measures in the full dataset (BESS (a) and SAC (b) total scores, and SCAT 5 symptom severity score (SCATSEV) (c) and total number of symptoms (SCATSYMP) (d))

Similarly, there were no associations in the normal RTP subset (BESS *p* = 0.58; SAC *p* = 0.937; SCATSEV *p* = 0.746; SCATSYMP *p* = 0.969) (*Supplementary figures 3a-d*) (Table 2). There were also no significant relationships in the long RTP subset (BESS *p* = 0.645; SAC *p* = 0.117; SCATSEV *p* = 0.465; SCATSYMP *p* = 0.578) (*Supplementary figures 4a-d*) (Table 2).

### Concussion recovery & severity across APOE genotypes

We used ANOVA tests to assess concussion recovery & severity by *APOE* genotype (ε3/ε3 vs. ε2 carriers, ε3/ε3 vs. ε4 carriers) (Table 3). Participants were divided based on whether they were a military or civilian participant. There were no differences between ε3/ε3 participants compared to ε2 carriers or ε4 carriers in either the military or civilian subgroups. There were also no significant differences when this analysis was repeated in only participants of European genetic ancestry (Table 3).

**Table 3.**
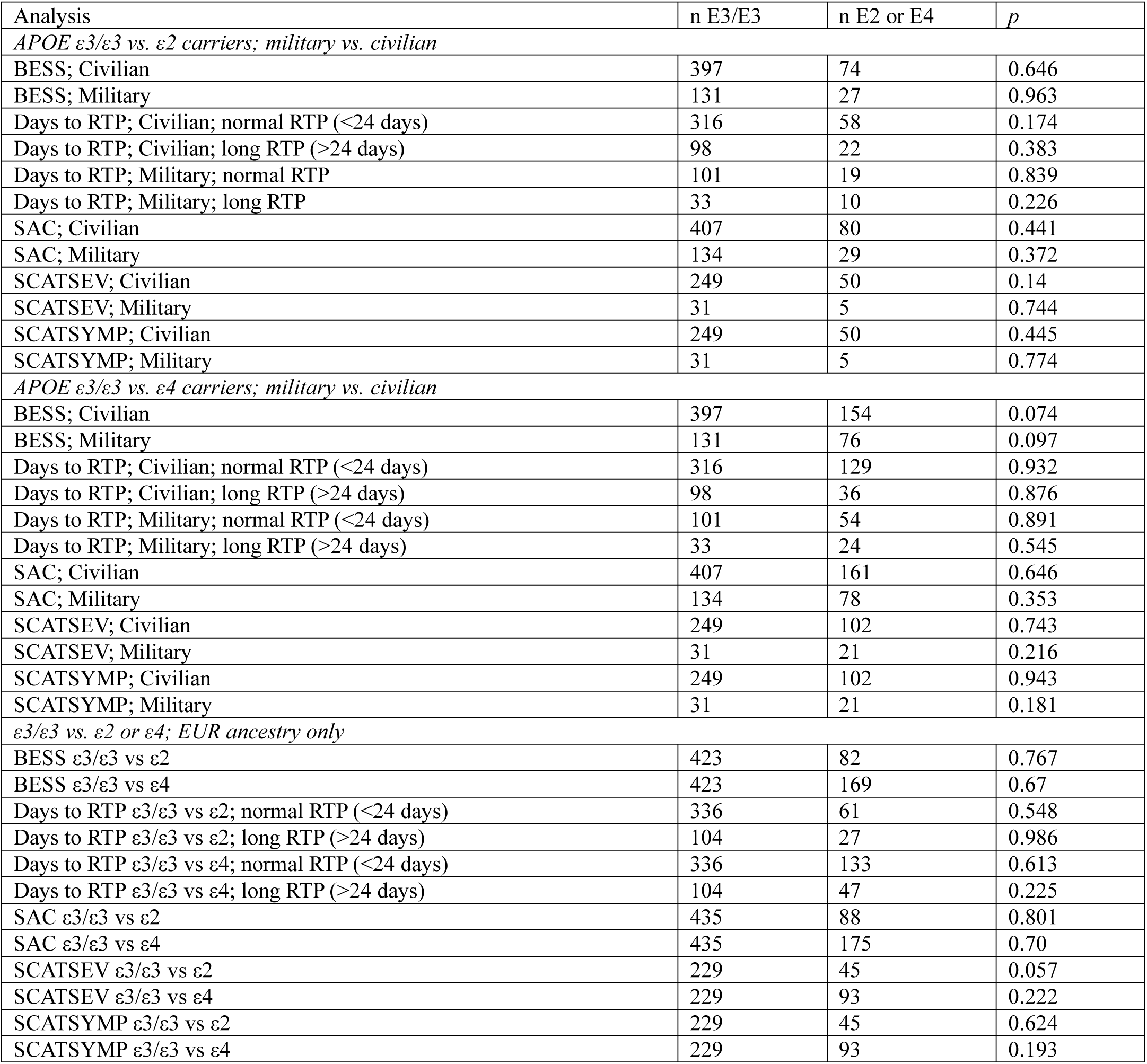
Results from ANOVAs of outcomes by *APOE* genotype (ε3/ε3 vs ε2 or ε4 carriers), participant type (military or civilian origin), and in participants of European genetic ancestry.

### Frequency of long vs. normal RTP by APOE genotype

We performed a chi-square analysis of RTP category by *APOE* genotype in military and civilian participants to test whether the frequency with which participants fell into the long RTP category was associated with *APOE* genotype. There were no significant findings in either civilian (ε3/ε3 vs ε2: χ2 (1, N = 494) = 0.534, *p* = 0.465; ε3/ε3 vs ε4: χ2 (1, N = 579) = 0.228, *p* = 0.633) or military (ε3/ε3 vs ε2: χ2 (1, N = 163) = 1.192, *p* = 0.275; ε3/ε3 vs ε4: χ2 (1, N = 212) = 0.946, *p* = 0.331) participants.

### Outcomes by PRS in European and African genetic ancestry

We used linear regressions to look for associations of PRS and outcome measures with participants divided based on genetic ancestry (African or European). There were no relationships in European participants (BESS *p* = 0.29; normal RTP *p* = 0.108; long RTP *p* = 0.86; SAC *p* = 0.673; SCATSEV *p* = 0.718; SCATSYMP *p* = .759) (Figures 4a-f). In individuals with African genetic ancestry, higher AD PRS was associated with lower SCAT total number of symptoms (SCATSYMP) (β = -1.796, SE = 0.794, 95% CI [-3.386, - 0.205], t-ratio = -2.260, *p* = 0.028) (Figure 5). There were no other significant differences (BESS *p* = 0.163; normal RTP *p* = 0.221; long RTP *p* = 0.446; SAC *p* = 0.715; SCATSEV *p* = 0.144) (*Supplementary figures 5a-e*).

**Fig. 4.**
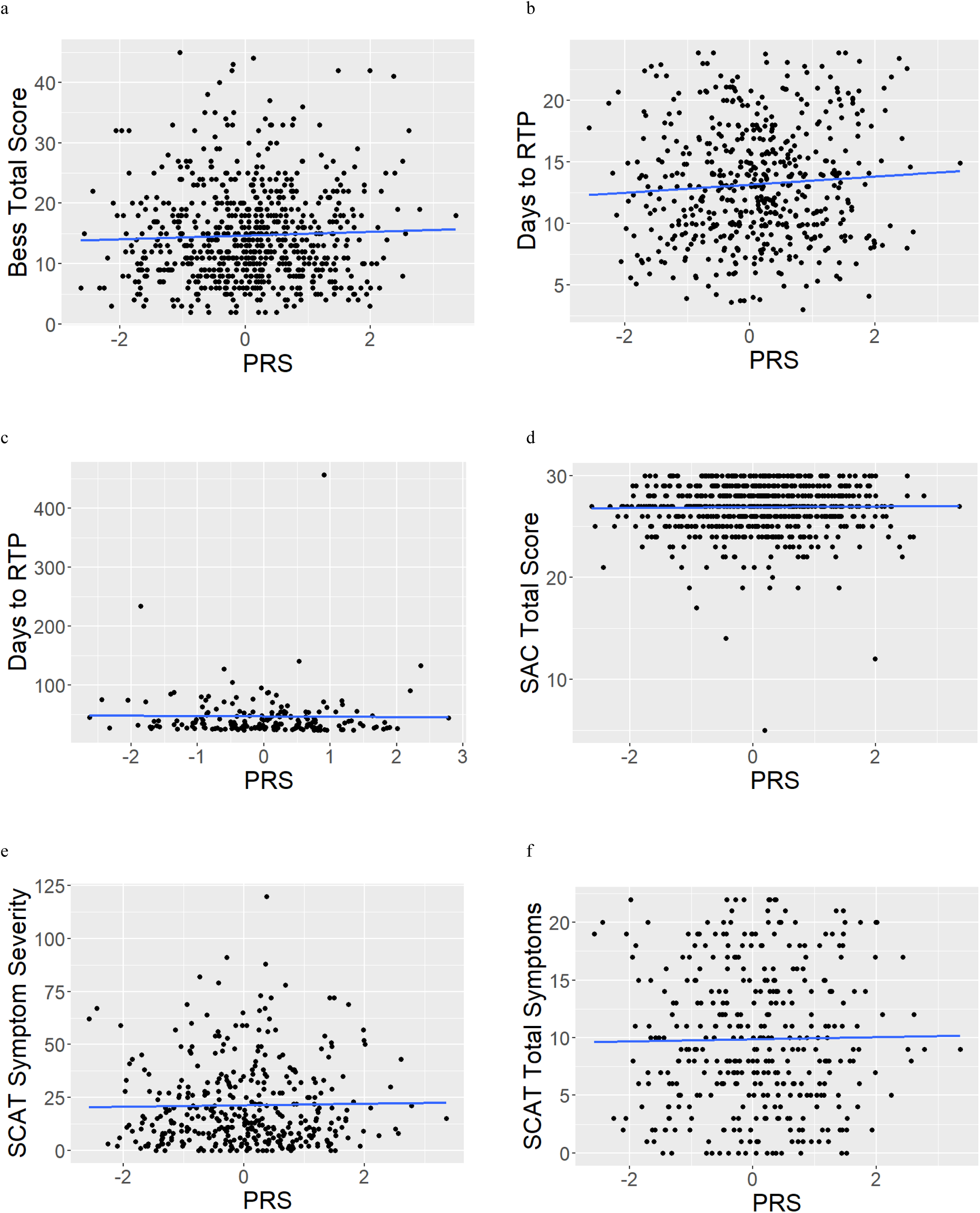
Outcome measures (BESS total score (a), days to RTP in normal (<24 days) subset (b), days to RTP in long (>24 days) subset (c), total score on SAC (d), SCAT symptom severity score (e), and SCAT total number of symptoms (f)) as a function of AD PRS in participants with European genetic ancestry.

**Fig. 5.**
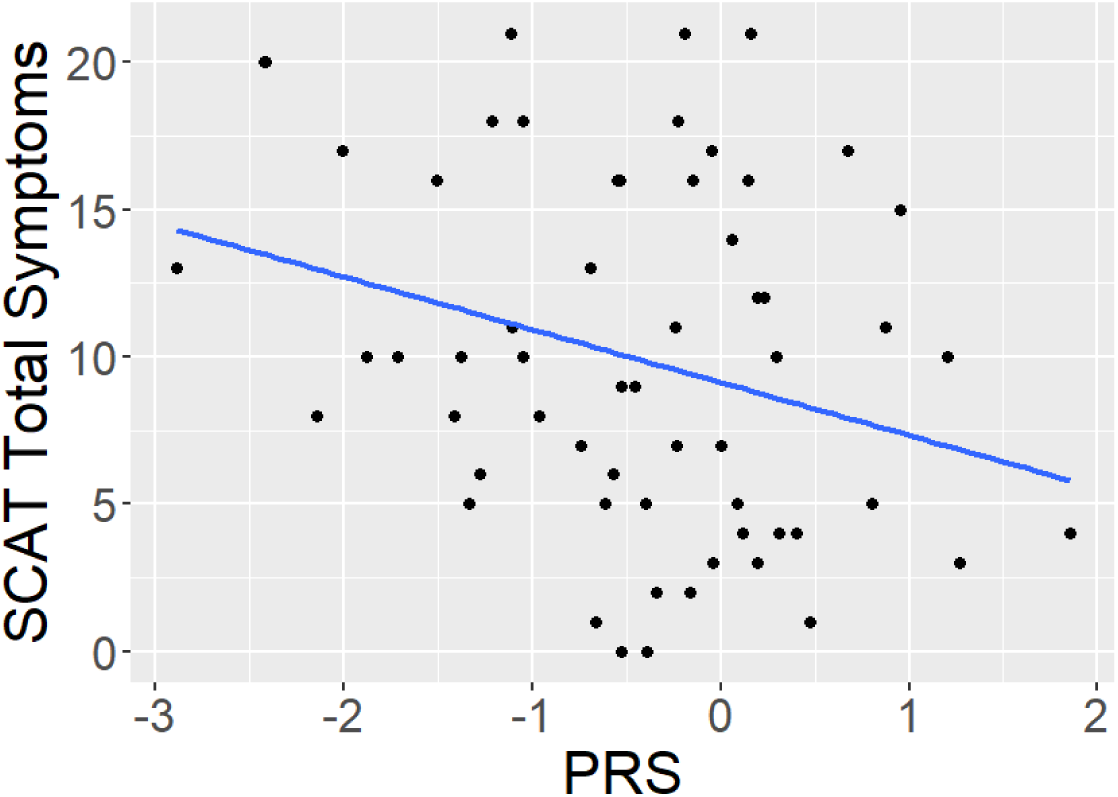
In individuals with African genetic ancestry, higher AD PRS was associated with lower SCAT total number of symptoms (SCATSYMP) (*p* = 0.0276)

## Discussion

Our preliminary results identified that an increase in AD PRS was associated with a slight increase to the injury to RTP interval in participants who took 24 days or less to RTP. However, as the remaining analyses and metrics were generally nonsignificant, our findings generally indicate that NCAA student athletes and Military Service Academy students with high genetic risk for AD are unlikely to experience worse concussions or poorer recovery than those with lower genetic risk.

Participants in the F LOC- group were 0.35 years (approximately 4.2 months) younger than participants in the M LOC- group. However, though this was statistically significant, a four-month age difference is unlikely to be clinically meaningful. Similarly, there was little evidence for an initial difference in injury or symptom severity between the groups. While the F LOC- group took longer to become asymptomatic than the M LOC- group, and the M LOC+ group demonstrated fewer symptoms on the SCAT5 than the F LOC- group, there was no consistent pattern of findings that suggested worse initial injury severity or recovery trajectories between the groups. Similarly, significant chi-square tests of both self-reported race/ethnicity and genetically derived ancestry are likely related to demographic differences in sports participation in the CARE Consortium, as described previously [64]. These significant tests are unlikely to represent clinically meaningful differences between groups on concussion severity and/or recovery.

Intriguingly, though the 21 *APOE* ε2/ε4 participants were excluded, 20 of these participants belonged to the M LOC- group while only one belonged to the F LOC- group. This is curious given that the *APOE* gene is located on chromosome 19 and is thus not expected to exhibit sex differences in prevalence. Furthermore, it was also surprising that we saw a significant association between AD PRS and concussion recovery, but no such association with *APOE*. This is especially surprising in the context of previous research showing *APOE* ε4 to be associated with poorer concussion outcomes [26–33]. This finding may suggest that continuous measures of genetic risk, such as PRS, are more sensitive measures to detect associations with concussion severity and/or recovery than categorical indicators of risk, like *APOE* genotype. However, replications are needed to further explore these possibilities.

The most interesting result from this analysis was our observation of a relationship between PRS and the injury to RTP interval in participants who took 24 days or less to return to play after an injury. In this group, higher AD PRS was associated with elevated recovery time. For every 1 standard deviation increase in PRS, the injury to RTP interval increased by 0.4121 days (9.89 hours). There was also a near-significant relationship between AD PRS and the injury to asymptomatic interval in the participants with <24 day RTP intervals, which supported this finding. However, the adjusted R^2^ value of the regression between AD PRS and the injury to RTP interval was 0.00585, indicating AD PRS only explains 0.585% of the variability in RTP. Furthermore, there was no relationship between AD PRS and either the injury to RTP or injury to asymptomatic intervals in the long (>24 days) RTP category. However, this group had fewer datapoints and numerous extremely long (100+ days) injury to RTP and/or asymptomatic intervals, both of which may partly contribute to this null finding. Additionally, upon subgroup analyses based on sex and LOC status of the injury to RTP interval within the normal RTP group, there were no significant relationships. Together, the results from this study do not provide compelling evidence for a consistent association between AD PRS and concussion severity or recovery.

Upon subgroup analyses based on genetic ancestry, we observed that higher PRS was associated with a lower SCAT total number of symptoms in individuals with African genetic ancestry, which was intriguing given that we would have expected higher AD genetic risk to be associated with more concussion symptoms. However, the R^2^ value was very small (multiple R^2^ = 0.081, adjusted R^2^ = 0.0651), indicating very little variance in the number of concussion symptoms can be attributed to PRS. Additionally, since the PRS was derived from European participants only, this finding in African genetic ancestry participants must be interpreted with significant caution.

Along these lines, an important limitation of our study is that the selected PRS may not have been an optimal choice. We chose this score because of the large sample size (over 400,000 subjects) used to generate it, but this population was entirely European [1]. The CARE dataset is not homogeneous (Table 1) [25], and prior reports have identified poor transferability of European scores into African ancestry populations [65, 66]. Though we performed some sub-group analyses based on genetic ancestry, these analyses should ideally be replicated using a PRS score more reflective of the demographics in CARE.

Another limitation is that the injury to RTP interval does not capture all dimensions of concussion recovery. Our method assumes that the interval is solely reflective of continued concussion symptomology, which may be an incorrect assumption in some cases. Recovery is a multi-factorial process, and many variables that can affect recovery trajectories were not considered here. For example, if an athlete suffered a simultaneous concussion and musculoskeletal injury, the injury to RTP interval could be influenced by the musculoskeletal injury, but we would be unable to distinguish this. Also, we did not consider psychological effects on recovery, but this is a critical future direction. Previous reports have identified relationships between concussion and psychological health in the form of anxiety/depression [67–72], particularly in individuals with persistent post-concussive symptoms [73–76]. Additionally, somatization is the biggest factor influencing concussion recovery [77]. As such, future studies would benefit by incorporating psychological test scores alongside physical recovery measures when assessing concussion recovery/severity in the context of genetic risk for neurodegenerative disease. An important future direction will be investigating the possibility of exploring PRS-associated neuroimaging changes in this cohort, such as white matter (WM) damage using diffusion tensor imaging (DTI). Reports from the CARE Consortium and other studies have found WM changes in concussed athletes/military servicemembers [78–84]. Using CARE data, one group compared DTI scans collected from 30 concussed football players within 48 hours of injury to 28 controls [80]. They found higher mean diffusivity (MD) in the concussed group [80], suggesting worsened myelin integrity. There were also correlations between axial diffusivity (AxD; indicative of axonal integrity), clinical outcomes (the Brief Symptom Inventory and the SCAT symptom severity score), and elevated recovery time [80]. The same group also studied 219 CARE participants with DTI scans collected at four time points: within 24 to 48 hours after injury, once asymptomatic, 7 days after RTP, and 6 months after injury [79]. Concussed athletes had higher MD both 24 to 48 hours after injury and once asymptomatic, and group differences in the corpus callosum were present at all time points [79]. Furthermore, elevated MD was also associated with increased recovery time [79]. These reports demonstrate the need to investigate WM damage in concussion as a function of AD genetic risk.

In general, there have been very few studies examining whether concussion-associated WM damage is exacerbated in individuals with high genetic risk for neurodegenerative disease, and this possibility warrants exploration. One study investigated the effect of *APOE* genotype on DTI metrics in concussion/mild traumatic brain injury and found limited evidence for altered fractional anisotropy (FA) in the cingulum of APOE ɛ4 carriers [85]. Another analyzed relationships between APOE ɛ4 genotype, blast exposure, and WM in military veterans, observing that ɛ4 carriers may be more vulnerable to WM abnormalities after blast exposure [86]. This evidence supports our hypothesis of a link between AD genetic risk and WM changes after concussion, as *APOE* ɛ4 is a known risk allele for both AD [34] and poorer prognosis after concussion [26–33]. However, additional evidence is lacking, and there has been little exploration into lesser-known AD risk genes using summary tools like PRS. These are important future directions to build on this project’s findings.

Finally, though we excluded repeated injuries from this analysis, investigating the effects of AD genetic risk on recovery after multiple injuries is an important next step. Though we did not identify consistent links between AD genetic risk and recovery/severity in the first injury exposure, it is possible that AD genetic risk may be more strongly associated with severity/recovery in the context of multiple injuries. We will investigate this possibility in future studies.

In conclusion, we observed an association of AD PRS with lengthened concussion recovery time based on one metric in collegiate athletes and military service academy students who were concussed during the CARE study. However, no additional analyses or metrics reached statistical significance, and it is therefore unlikely that AD genetic risk is strongly associated with concussion severity or recovery. Future studies should attempt to replicate these analyses in larger samples with longer follow-up using PRS calculated from multiple/diverse populations to clarify and add to these findings. Additionally, incorporating multiple modulators and metrics of injury recovery, including psychological health, coincident non-concussion injuries, and neuroimaging metrics such as diffusion tensor imaging, will be of significant value.

## Data Availability

Those outside the consortium can access de-identified CARE data via FITBIR, the Federal Interagency Traumatic Brain Injury Research database.

https://fitbir.nih.gov/

## Author Contributions

KMD was responsible for data analysis and drafting the manuscript. KN contributed to study conceptualization and design, data analysis, and revision of the manuscript. TWM, YCW, BCM, SLR and AJS contributed to study conception and revision of the manuscript. MM, SB, PP, MB, JM, KG, CG, JG, SD, SR, SS, KC, MH, DC, GM, and JJ contributed to data collection and revision of the manuscript. All authors read and approved the final version of the manuscript.

## Acknowledgements

The authors thank Gillian Ising and Jody Harland for their assistance.

