## Supplemental information for "Association of Alzheimer’s disease polygenic risk score with concussion severity and recovery metrics"

**Supplementary fig. 1.** Sensitivity analysis of missing SNP data handling methods. In (a) and (b), linear regressions to test the relationship between RTP and AD PRS were performed on the full 931 participants using two methods for handling missing SNP data: the ignoring method (PRS value of missing SNP is replaced with 0) (a) and the mean substitution method (PRS value of missing SNP is replaced with the mean score for that SNP from all other participants) (b). For the ignoring method, there was no significant relationship ($p = 0.8152$). The relationship approached significance using the mean substitution method ($p = 0.05314$). These tests were repeated using a subsample of participants with outliers removed by R (c and d). There were no significant relationships using the ignoring method ($p = 0.8605$) (c) or the mean substitution method ($p = 0.7352$) (d).

a

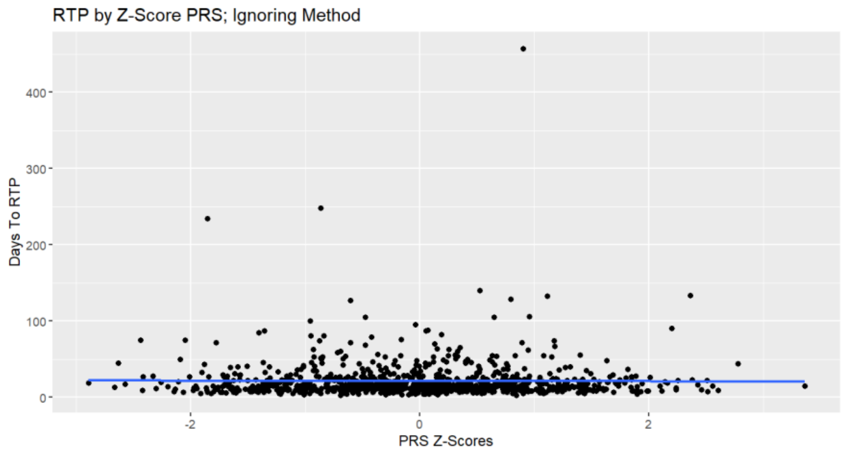


b

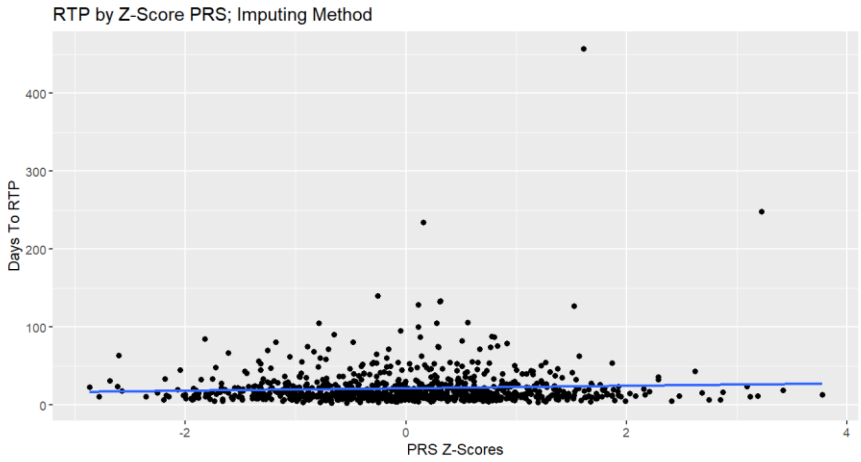

c

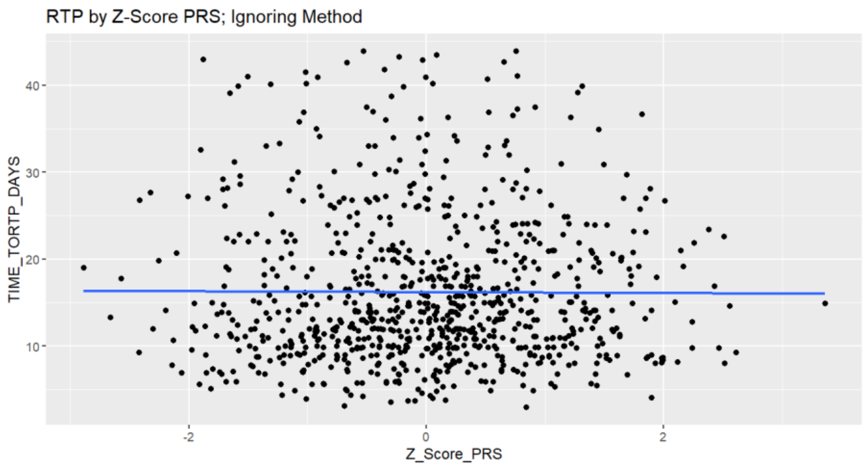

RTP by Z-Score PRS; Ignoring Method
TIME_TORTP_DAYS
40
30
20
10
-2
0
2
Z_Score_PRS


d

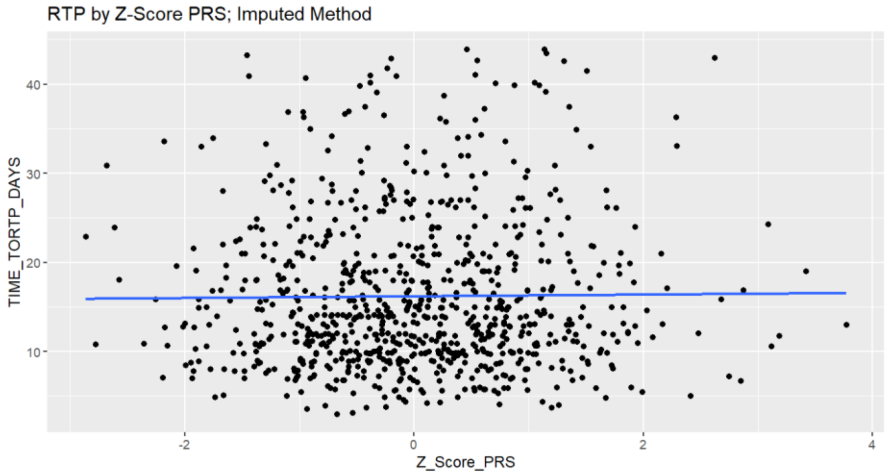

RTP by Z-Score PRS; Imputed Method
TIME_TORTP_DAYS
40
30
20
10
-2
0
2
4
Z_Score_PRS

**Supplementary fig. 2.** AD PRS & normal RTP interval in F LOC+ (a), F LOC- (b), M LOC+ (c), and M LOC- (d) participant subgroups.

a

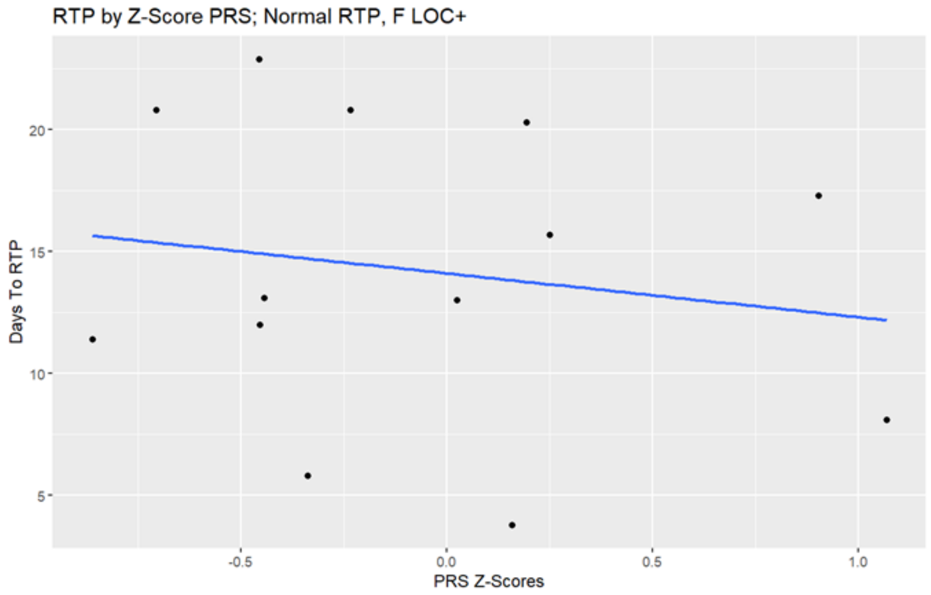


b

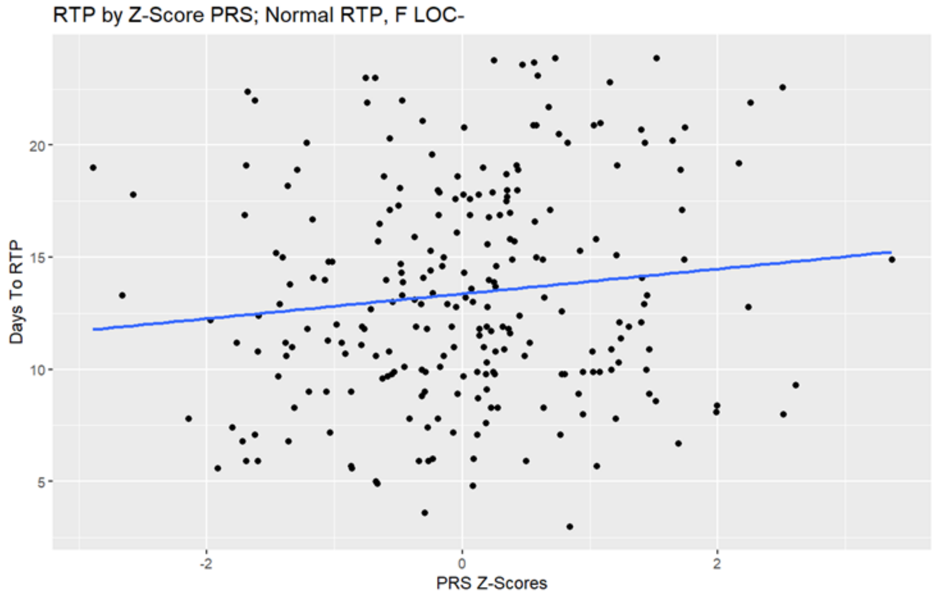

c

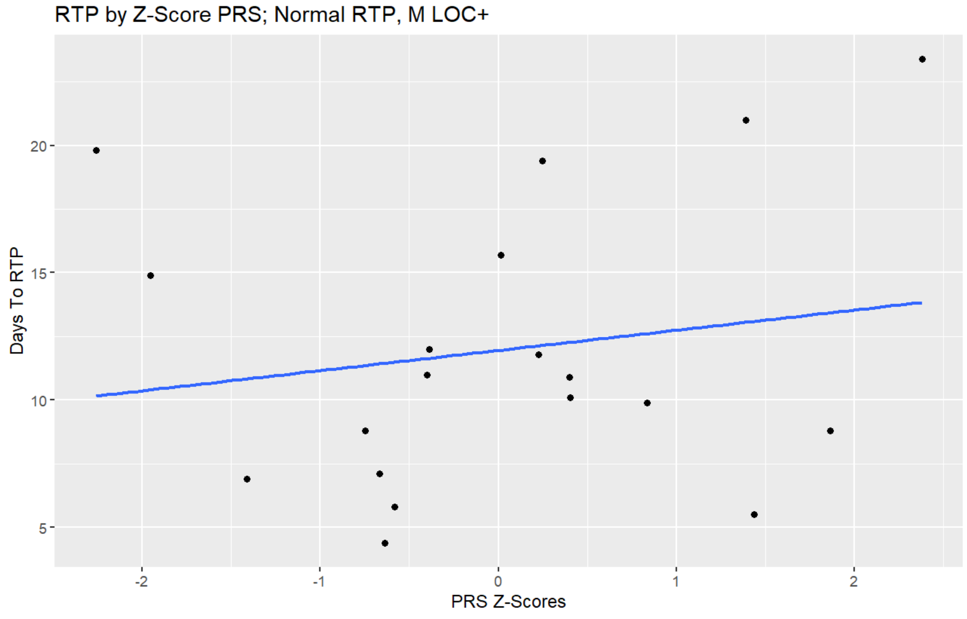
RTP by Z-Score PRS; Normal RTP, M LOC+
Days To RTP
5
10
15
20
-2
-1
0
1
2
PRS Z-Scores

d

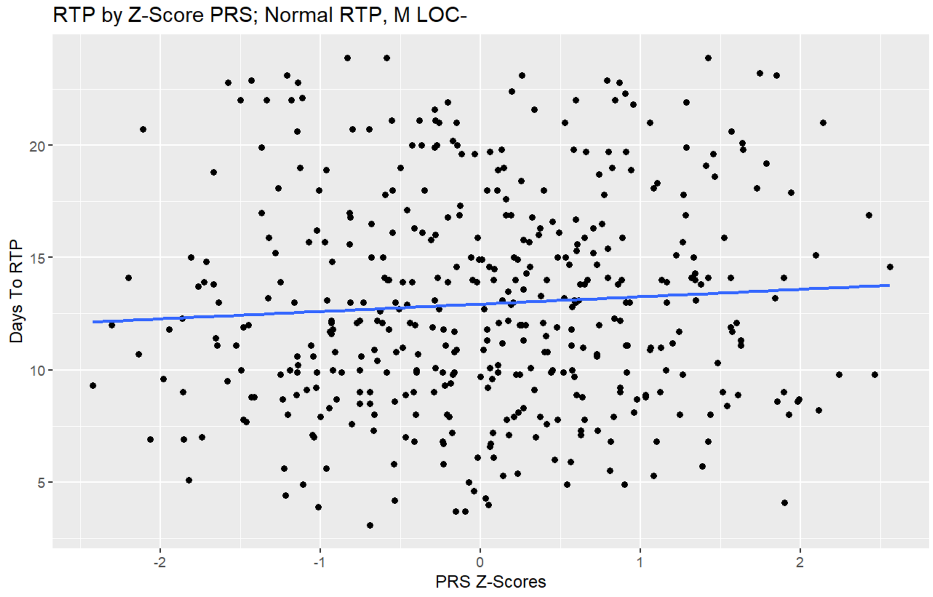
RTP by Z-Score PRS; Normal RTP, M LOC-
Days To RTP
5
10
15
20
-2
-1
0
1
2
PRS Z-Scores

**Supplementary fig. 3.** AD PRS & total scores on BESS ($p = 0.58$) (a), total scores on SAC ($p = 0.937$) (b), SCAT symptom severity scores (SCATSEV; $p = 0.746$) (c), and SCAT total number of symptom scores (SCATSYMP; $p = 0.969$) (d) in normal RTP (<24 days) data subset.

a

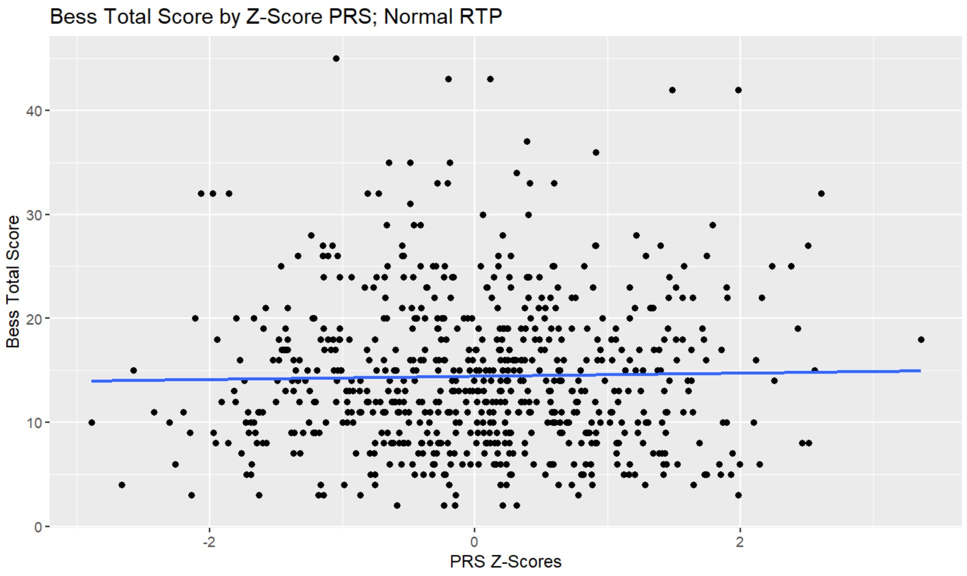


b

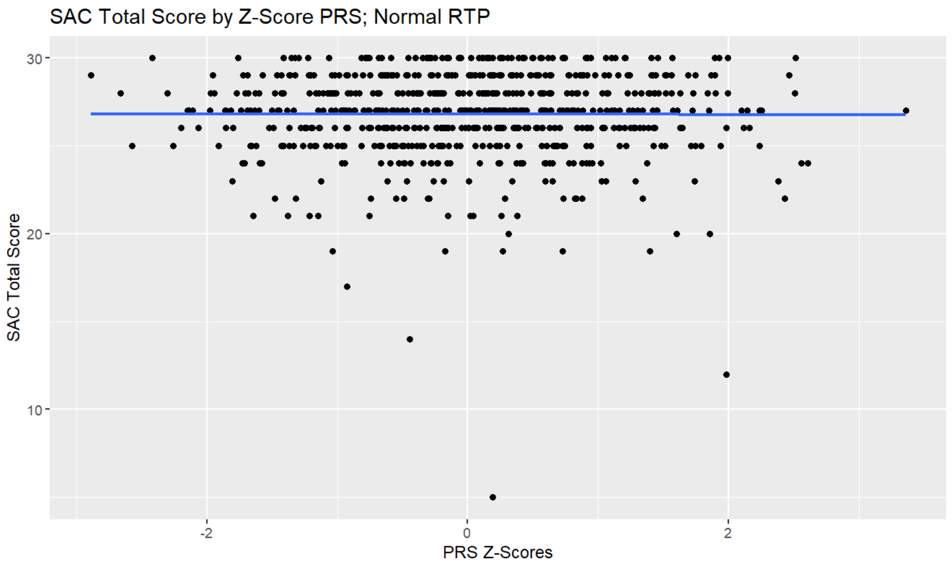

c

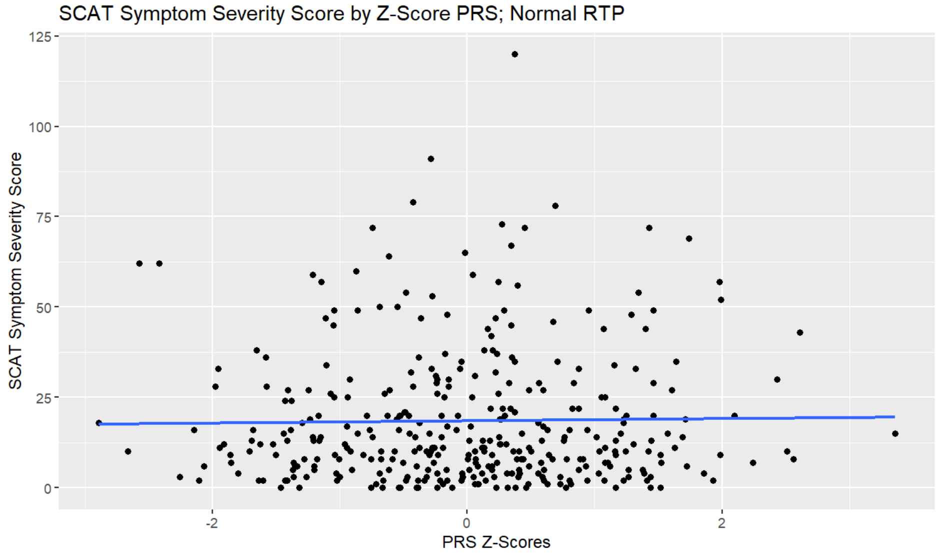
SCAT Symptom Severity Score by Z-Score PRS; Normal RTP
SCAT Symptom Severity Score
125
100
75
50
25
0
-2
0
2
PRS Z-Scores

d

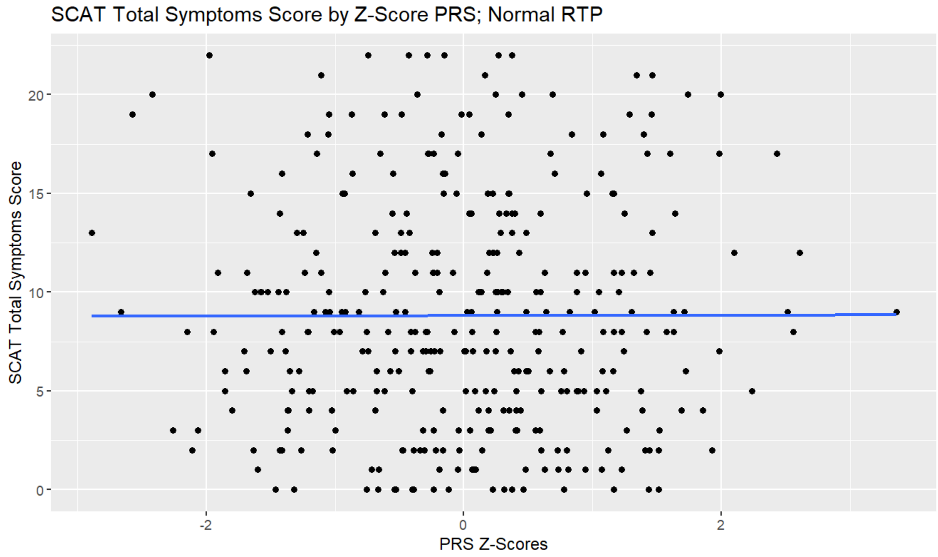
SCAT Total Symptoms Score by Z-Score PRS; Normal RTP
SCAT Total Symptoms Score
20
15
10
5
0
-2
0
2
PRS Z-Scores

**Supplementary fig. 4.** AD PRS & total scores on BESS ($p = 0.645$) (a), total scores on SAC ($p = 0.117$) (b), SCAT symptom severity scores (SCATSEV; $p = 0.465$) (c), and SCAT total number of symptom scores (SCATSYMP; $p = 0.578$) (d) in long RTP (>24 days) data subset.

a

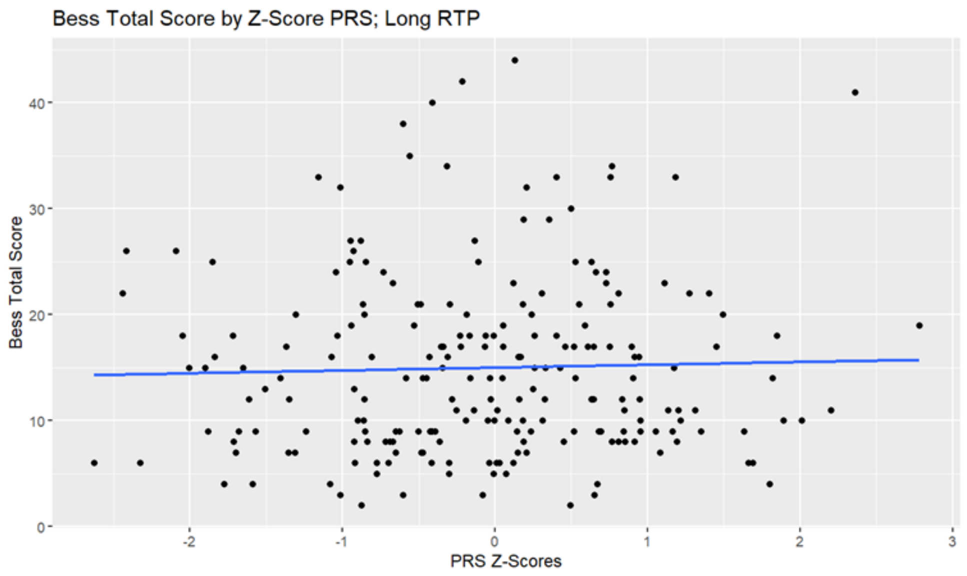


b

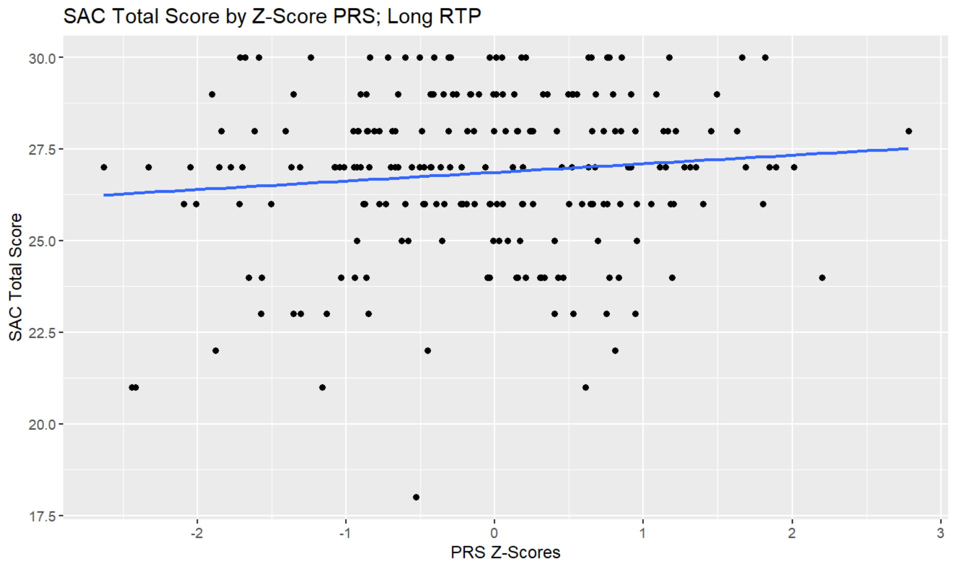

c

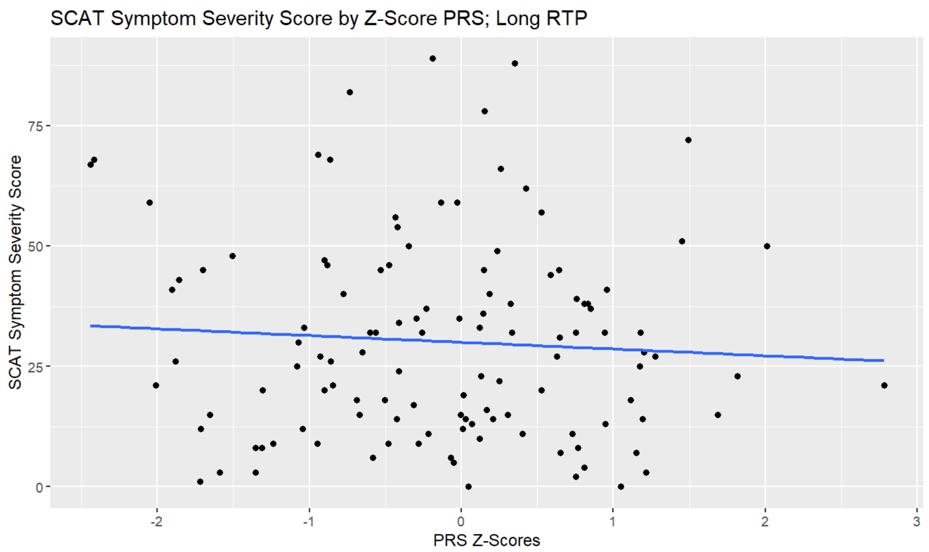
SCAT Symptom Severity Score by Z-Score PRS; Long RTP
SCAT Symptom Severity Score
0
25
50
75
-2
-1
0
1
2
3
PRS Z-Scores

d

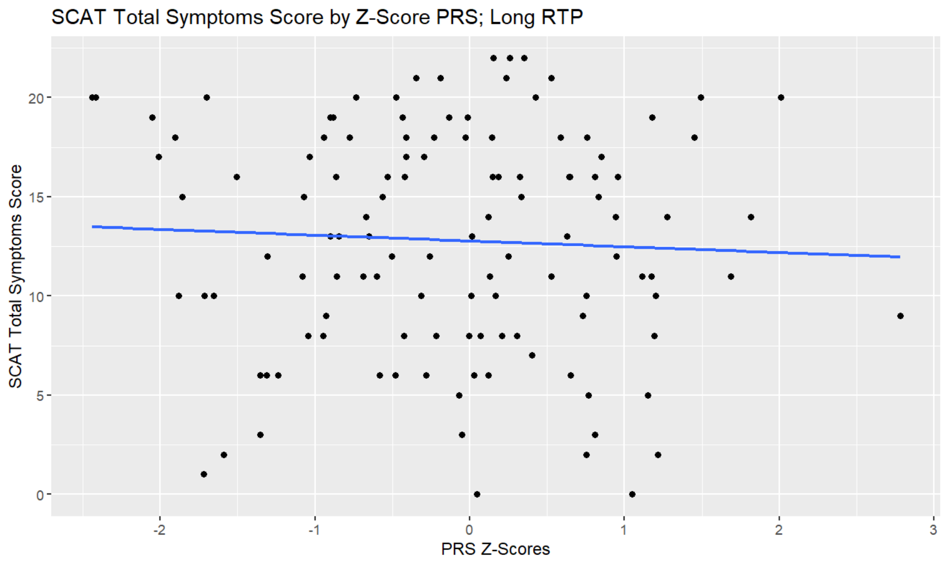
SCAT Total Symptoms Score by Z-Score PRS; Long RTP
SCAT Total Symptoms Score
0
5
10
15
20
-2
-1
0
1
2
3
PRS Z-Scores

**Supplementary fig. 5**. AD PRS and total scores on BESS (*p* = 0.163) (a), days to normal RTP (*p* = 0.221) (b), days to long RTP (*p* = 0.446) (c), total scores on SAC (*p* = 0.715) (d), and SCAT symptom severity scores (SCATSEV; *p* = 0.144) (e) in participants of African genetic ancestry.

a

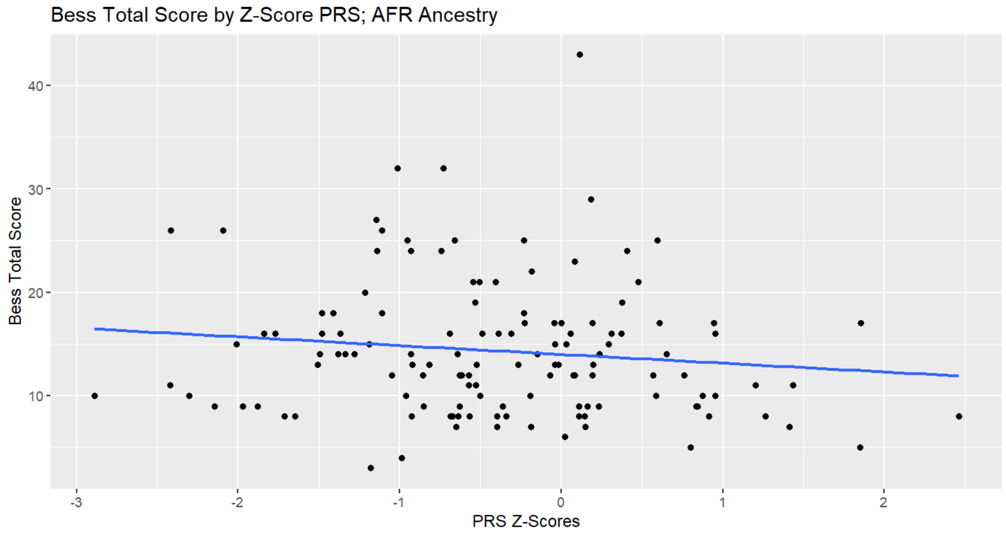


b

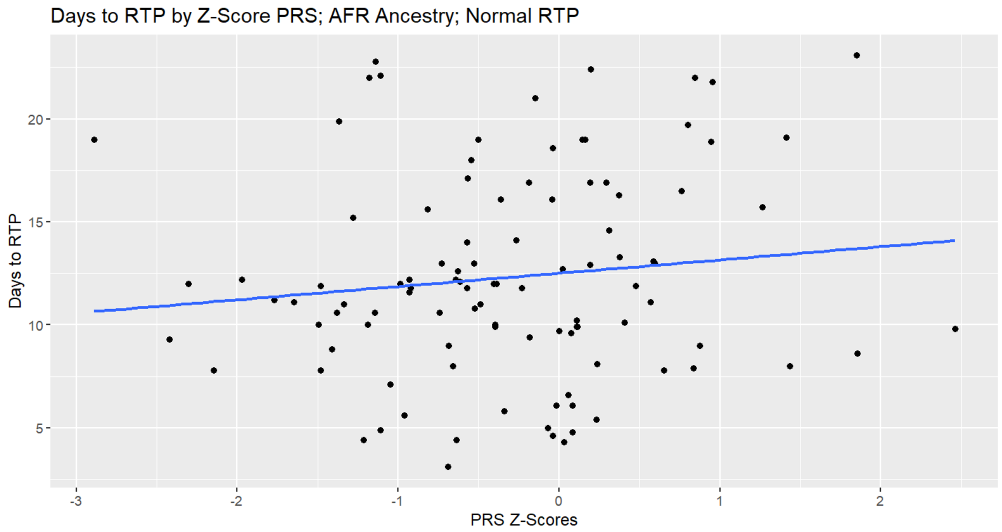

c

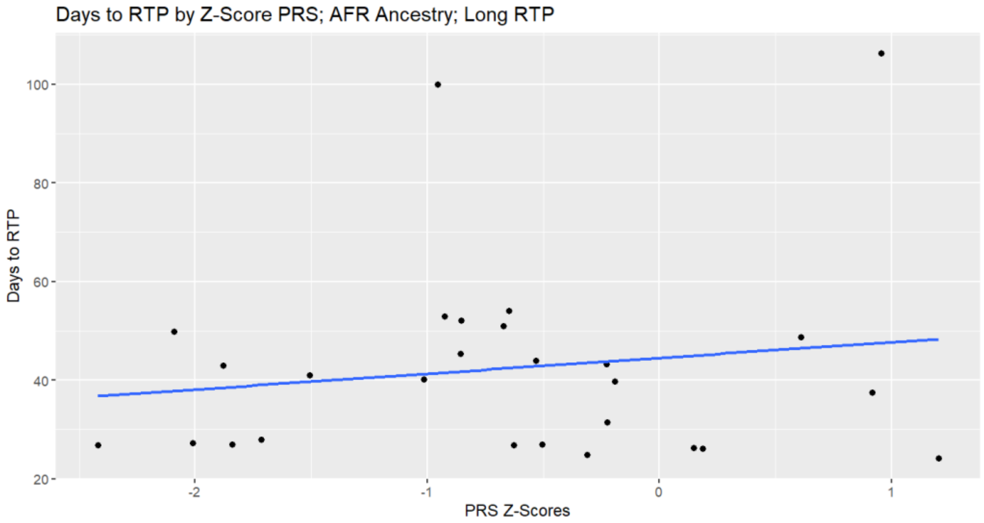
Days to RTP by Z-Score PRS; AFR Ancestry; Long RTP
Days to RTP
100
80
60
40
20
-2
-1
0
1
PRS Z-Scores

d

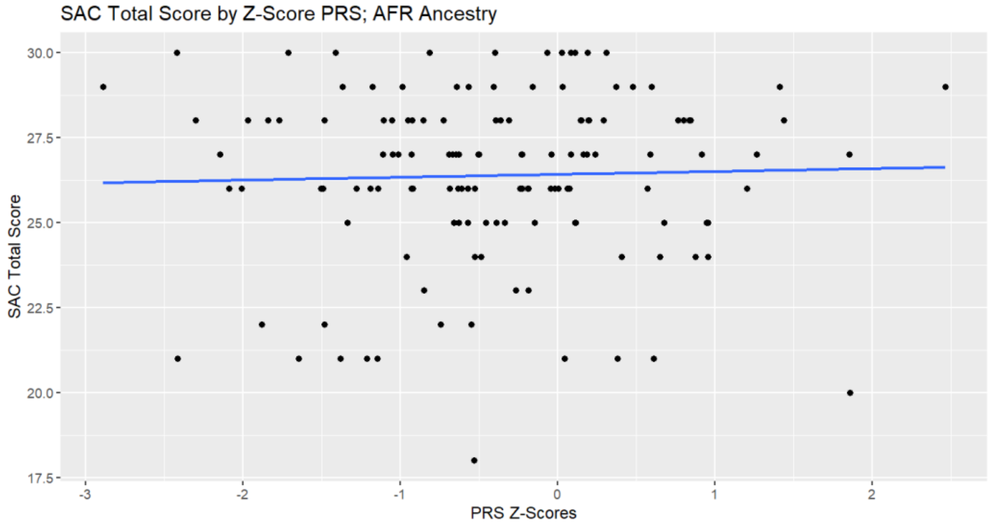
SAC Total Score by Z-Score PRS; AFR Ancestry
SAC Total Score
30.0
27.5
25.0
22.5
20.0
17.5
-3
-2
-1
0
1
2
PRS Z-Scores

e

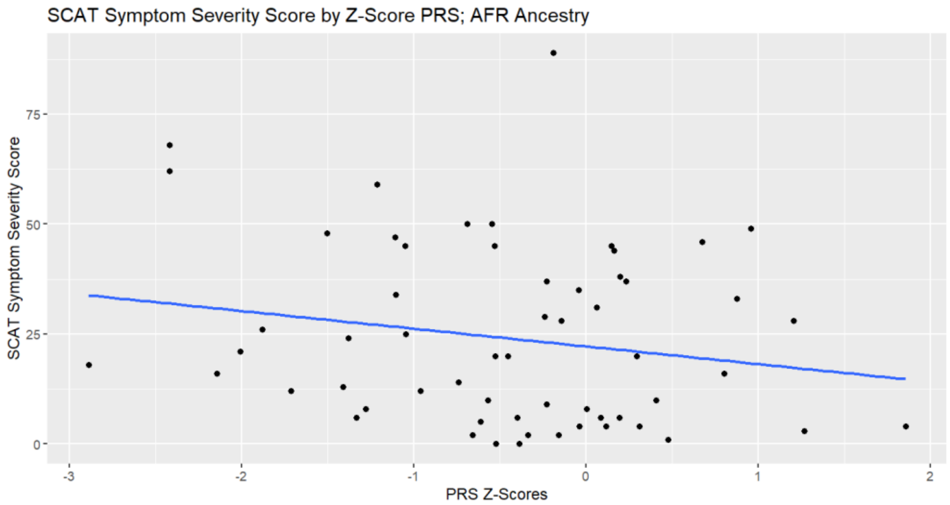

SCAT Symptom Severity Score by Z-Score PRS; AFR Ancestry
SCAT Symptom Severity Score
75
50
25
0
-3
-2
-1
0
1
2
PRS Z-Scores
